# Altered immunity to microbiota, B cell activation and depleted γδ / resident memory T cells in colorectal cancer

**DOI:** 10.1101/2021.02.15.21251750

**Authors:** Alistair Noble, Edward T Pring, Lydia Durant, Ripple Man, Stella M Dilke, Lesley Hoyles, Steve A James, Simon R Carding, John T Jenkins, Stella C Knight

## Abstract

We sought methods of rectifying microbiota:immune dysregulation as a route to prophylaxis and improved immunotherapy of colorectal cancer (CRC). CRC develops in gut epithelium, accompanied by low level inflammatory signaling, intestinal microbial dysbiosis and immune dysfunction. We examined populations of intraepithelial lymphocytes in non-affected colonic mucosa of CRC and healthy donors and circulating immune memory to commensal bacterial species and yeasts. Colonic tissue in CRC was significantly depleted of γδ T cells and resident memory T cells, populations with a regulatory CD39-expressing phenotype. T cell memory responses to a panel of commensals were distinct in CRC, while B cell memory responses to several bacteria/yeast were significantly increased, accompanied by increased proportions of effector memory B cells, transitional B cells and plasmablasts in blood. IgA responses to mucosal microbes were unchanged. Our data describe a novel immune signature with similarities to and differences from that of inflammatory bowel disease. They implicate B cell dysregulation as a potential contributor to parainflammation and identify pathways of weakened barrier function and tumor surveillance in CRC-susceptible individuals.

## Introduction

Colorectal cancer (CRC) arises from a complex interplay between genetic, inflammatory, environmental and microbiome-related factors. CRC usually arises from adenomatous polyps derived from intestinal stem cells and can be caused by genetic mutations, including lesions in the adenomatous polyposis coli (APC) gene (associated with familial adenomatous polyposis) or mismatch repair genes (e.g. Lynch syndrome). Globally, in 2012 there were an estimated 1.4 million new cases and almost 700,000 deaths attributable to CRC. Around 2% of cases arise in those with inflammatory bowel disease (IBD) and 15% of IBD patients die from CRC^1^. Cancer is associated with low-grade or parainflammation^2^ and sustained raised levels of circulating pro-inflammatory cytokines, e.g. TNF-α, are observed in CRC patients^3^. Therefore, it appears that CRC develops in an environment of immune misdirection, in which unidentified antigens drive parainflammation while cell-mediated immunity to tumor-associated antigens fails.

The composition of the microbiota and the phenotype of dendritic cells (DC) are altered in CRC^4,5,6^. The intestinal microbiota constantly interacts with the immune system at the mucosal interface, with DC continually sampling contents of the lumen and presenting microbial and food antigens to the lymphocytes situated in mucosal-associated lymphoid tissue. In mouse models, colitis-associated CRC is prevented by T-bet expression in DC which is T-cell-dependent^7^. T-bet^+^ DC are crucial to initiation of cell-mediated immune responses dominated by CD8^+^ T cells. CD8^+^ T cells are a prominent tissue-resident population within the gut mucosa, being abundant in both the epithelium and lamina propria. γδ T cells are another prominent T cell population making up intraepithelial lymphocytes (IEL), expressing a natural tissue-resident phenotype, and are crucial to tumor surveillance due to their ability to kill tumor cells irrespective of conventional MHC class I restriction and dependent on their unconventional receptor specificities^8^. γδ T cells can respond to microbes via Toll-like receptors or via epithelial cells, to maintain barrier function and tissue repair^9^.

Resident memory T cells (Trm), which are non-circulating CD103- and CD69-expressing cells, can reside in both mucosal tissue and within tumors. High numbers of Trm within tumor tissue are associated with a good prognosis^10^. Tumor Trm cells are believed to strongly adhere to their target tumor cells via CD103 binding to its ligand E-cadherin, expressed on epithelial cells^11^. Precursors of Trm may traffic from blood to the inflamed tumor environment and develop Trm-like characteristics, with antigen specificities that may include tumor-associated markers or antigens originating from the tumor-associated microbiota. In IBD, loss of γδ and Trm populations is associated with abnormal B cell responses to the microbiota, especially in Crohn’s disease^12^. This suggests that poor cell-mediated immunity results in increased microbial translocation into gut tissues and uncontrolled B cell responses. Since a disturbed microbiota is associated with both IBD and CRC, and IEL are closely associated with, and respond to, microbial antigens from the microbiota, we set out to test the hypothesis that γδ or Trm populations in the non-lesional mucosa of CRC-susceptible individuals are involved in development of parainflammation and/or tumorigenesis. Ours is the first study to associate depletion of these immune populations with CRC and to identify B cell dysfunction as a potential contributor to parainflammation.

Our data implicate a dysfunctional relationship between the immune system and microbiota in the loss of tissue-resident populations which are critical in tumor immune surveillance. The results identify novel immune markers in tissue and blood that associate with susceptibility to CRC.

## Materials and Methods

### Study Design

Donors (age 16-80 years) were recruited from St Mark’s Hospital, the UK national bowel hospital, and included patients undergoing surgery for CRC and healthy donors undergoing investigative endoscopy. Patients were recruited between June 2018 and October 2019; no data were excluded at the end of the study. Clinical and demographic patient characteristics are shown in Table 1. Surgical tissue from the left colon and/or 20 ml peripheral blood was obtained from participants. Surgery patients were treated with antibiotics (neomycin) and mechanical bowel preparation (laxatives), as part of routine bowel preparation within the 24-hour period prior to surgery. Healthy gut tissue donors received mechanical bowel preparation only. Additional healthy blood donors were recruited from hospital staff and visitors. Ethical approval was obtained from the Health Research Authority UK and London-Harrow Research Ethics Committee (study ref 17/LO/1636). Written informed consent was received from participants prior to inclusion in the study.

**Table 1.**
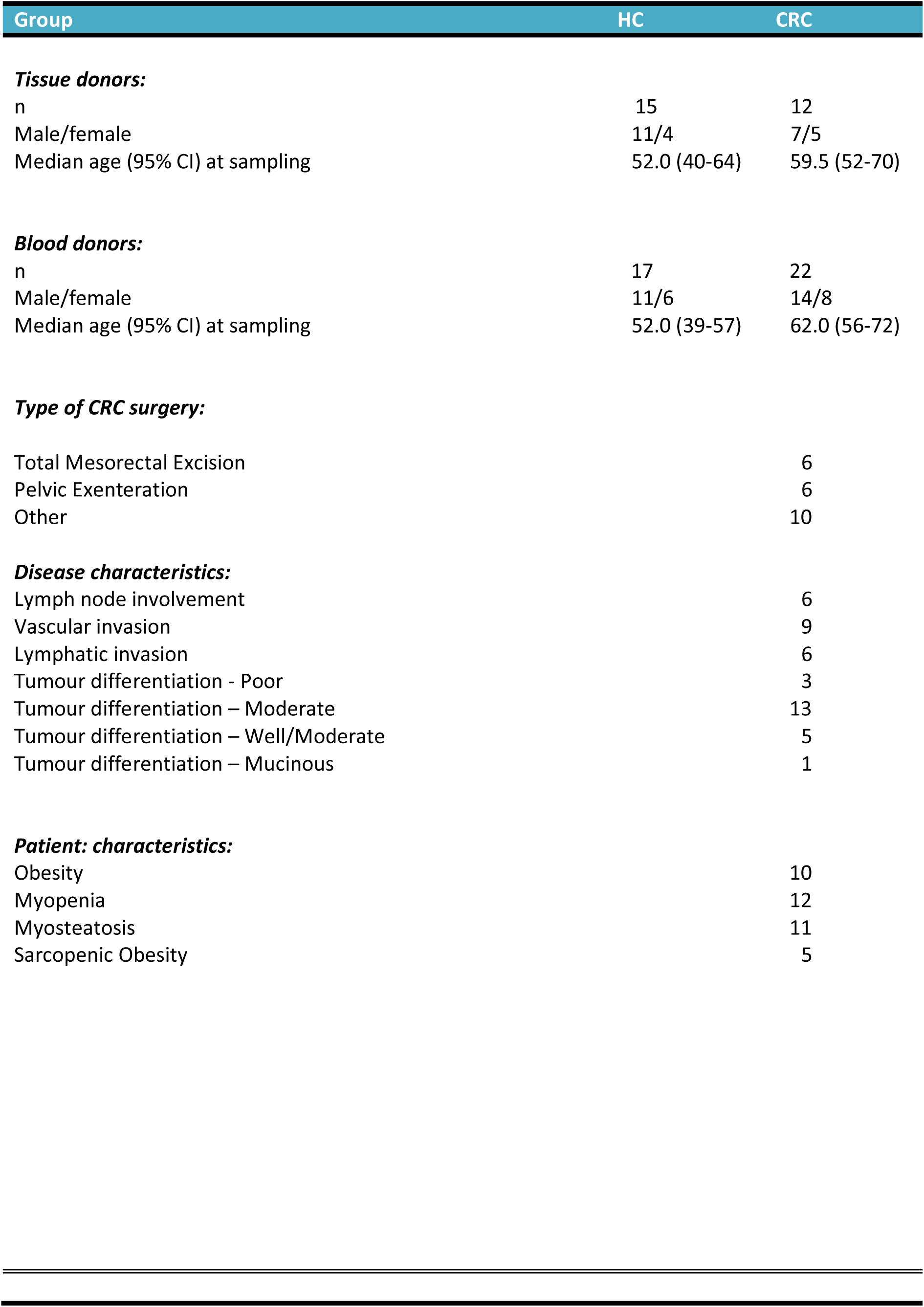
Clinical characteristics of CRC and healthy control (HC) participants.

### Colonic intraepithelial lymphocytes (IEL), lamina propria lymphocytes (LPL) and intraepithelial microbe (IEM) isolation/characterization

For some CRC patients, 5 left colon mucosal biopsies (10 mg tissue each) were obtained from macroscopically non-lesional tissue sites of CRC surgical tissue post resection. For healthy control tissue donors, five 10 mg left colon biopsies were obtained from macroscopically non-lesional tissue sites during routine colonoscopy. IEL and IEM were released from biopsies using DTT/EDTA and harvested by centrifugation at 300 *g* (5 min). IEM were obtained by centrifugation of resulting supernatants at 4500 *g* (20 min). LPL were obtained by digestion with collagenase of remaining tissue. All cells were washed, then phenotyped and counted by flow cytometry. Cells were first stained for viability using LIVE/DEAD Fixable-near-IR stain (ThermoFisher) before addition of surface-staining antibodies in fetal calf serum. In some cases, cells were then fixed/permeabilized for intranuclear staining using the Foxp3 buffer set (ThermoFisher, as instructions). Cells were stained with antibodies to γδ TcR, CD4, CD8, CD103, CD69, CD39 and CD73 (supplementary Table 1). All samples were acquired on a BD Biosciences FACS Canto II flow cytometer and data analyzed by FlowJo software (Tree Star), with volumetric sampling determined using Perfect-Count microspheres™ (Cytognos, S.L).

### Commensal-specific T and B cell memory proliferative responses

Commensal species were isolated from the cecum of healthy donors^13,14^. Strains were grown anaerobically in Hungate tubes containing Wilkins-Chalgren broth (37°C for 24 h). Aliquots (1 ml) were centrifuged (13,000 rpm for 10 min), supernatants removed and cell pellets snap-frozen on dry ice before storage at −80°C. Yeast strains were isolated from a stool sample collected from a healthy young infant donor. Strains were grown statically in yeast-extract peptone (YPD) broth (37°C for 72 h). Aliquots (1 ml) were centrifuged (1000 *g* for 15 min), supernatants removed and cells resuspended in an equal volume of PBS. Yeast cells were killed by heat treatment (70°C for 3 h) and heat-treated cell suspensions were stored at 4 °C until required. PBMC were obtained over Ficoll gradients and labelled with CellTrace Violet™ (1 µM, Life Technologies) according to manufacturer’s instructions, then cultured at 4×10^6^/ml in XVIVO15 serum-free medium (Lonza, +50µg/ml gentamycin (Sigma) and penicillin/streptomycin (ThermoFisher, 1/100)). Killed cells (2×10^5^) from 9 bacterial species and two fungal species (listed in Fig 4) were added to 0.2 ml cultures and microbe-specific CD4^+^/CD8^+^ T cell and B cell proliferative responses were determined after 7 days culture. Cultured cells were analyzed by staining with LIVE/DEAD stain and CD4/CD8/CD19 antibodies (supplementary Table 1).

### Measurement of antibody responses

IEM were labelled with SYBR Green DNA stain (ThermoFisher, 1/100,000), anti-IgA-APC/anti-IgG-APC/Cy7 and analyzed by flow cytometry to determine proportions (%) of bacteria coated with antibodies in the gut. Intact microbes were gated according to SYBR Green staining.

### Statistical analysis

GraphPad Prism 9 software (GraphPad, San Diego, CA) was used to plot and analyze the data, using Mann Whitney non-parametric tests. For proliferation data Kruskal-Wallis ANOVA was used, with correction for multiple comparisons applied using the Benjamini, Krieger & Yekutieli method. P values less than 0.05 were considered significant and indicated by: *, p<0.05; **, p<0.01; ***, p<0.001.

## Results

### γδ T cells with residence markers and CD8^+^ and CD4^+^ Trm are deficient in CRC mucosa

γδ T cells and resident memory T cells were identified in both IEL and LPL from healthy colonic endoscopic biopsies using the extracellular markers CD103 and CD69 and the intranuclear transcription factor Runx3^15^ (Figure 1). IEL consisted of γδ T cells and CD8^+^ T cells, the majority of which expressed both CD103 and CD69 and were therefore classed as a tissue resident phenotype. Consistent with our previous study^12^, CD103^+^ T cells preferentially expressed CD39, a regulatory T cell-associated ectonucleotidase not present on circulating T cells. Indeed, Trm express both pro-inflammatory and regulatory features^12^. Among LPL, γδ T cells were rare and large numbers of CD4 and CD8 T cells were present, lower proportions of which expressed CD103 and CD39 compared to IEL although CD69 was still present (Figure 1a). Runx3, the transcription factor that drives Trm differentiation^15^, was absent in LPL but could be seen at high levels in some γδ T cells and at lower levels in most (50-80%) CD8^+^ IEL (Figure 1b).

**Figure 1.**
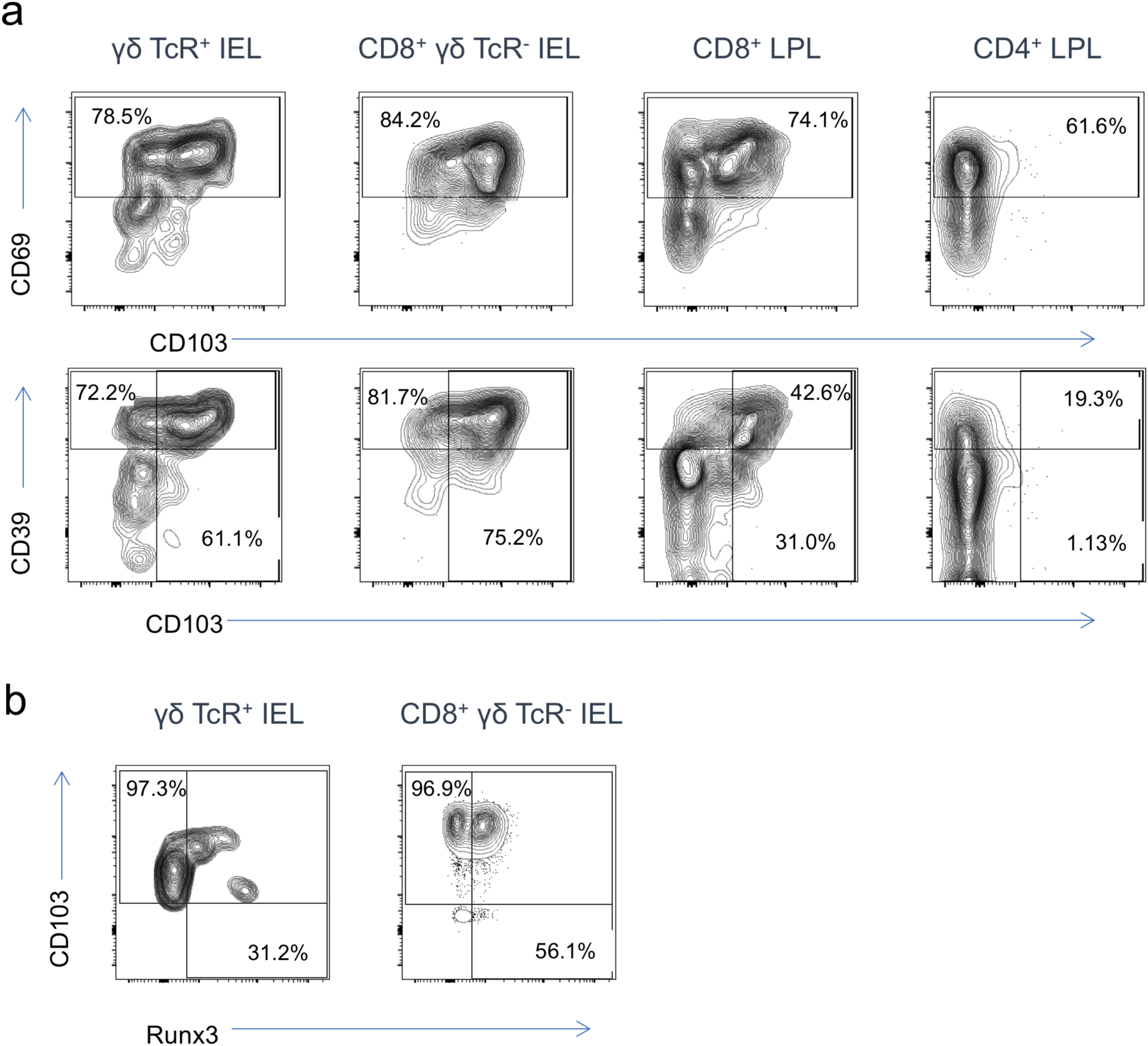
Colonic γδ T cells and Trm expressing the CD103 surface marker and Runx3 transcription factor preferentially express CD39. A: CD8 T-cell and γδ T-cell populations were identified in IEL fractions (left panels) and CD8 and CD4 T cells were identified in LPL fractions (right panels). Cells were stained for CD69 and CD103 surface Trm markers (upper panels) or CD103 and CD39 (lower panels). B: IEL populations were permeabilized for intranuclear detection of Runx3 transcription factor; no Runx3 was seen in LPL (not shown). All data are from healthy donors and representative of three independent experiments.

We then analyzed the presence of γδ T cells and Trm in non-affected colonic mucosa from CRC patients, to test the hypothesis that there is an underlying and detectable susceptibility to tumorigenesis in colonic tissue. Surgical tissue was used as a source of mucosal biopsies, taken by forceps in a manner as comparable as possible to biopsy via endoscopy. Numbers and proportions of tissue-resident mucosal T cells are shown in Figure 2, using CD103 as the defining marker. Both total numbers of T cells and percentages of total cells (which included epithelial cells) were dramatically reduced in CRC biopsies compared to those from healthy donors. This was the case for all CD103-expressing cells including γδ T cells in IEL, CD8^+^ Trm in IEL and LPL, and CD4^+^ Trm in LPL, proportions of which in CRC samples represented on average 14%, 15%, 9.0% and 5.5% of those in healthy donors, respectively. Furthermore, the proportion of cells expressing CD103 within each subset was significantly reduced in CRC patients (with the exception of CD8^+^ Trm in LPL which did not reach statistical significance) (Figure 2a-d). Thus, CRC tissue exhibits a profound and selective depletion of tissue resident lymphocytes, which is comparable, though more extreme, than that we have described in IBD^12^. Only one of the CRC patients had been diagnosed with IBD (Crohn’s disease). The depletion could not be accounted for by a concentration of T cells in tumor tissue itself, as this contained similar numbers of T cells to surrounding tissue (unpublished observation). Furthermore, depletion of the same populations was seen in pre-cancerous individuals with familial adenomatous polyposis (unpublished data), indicating that immune dysfunction precedes tumorigenesis.

**Figure 2.**
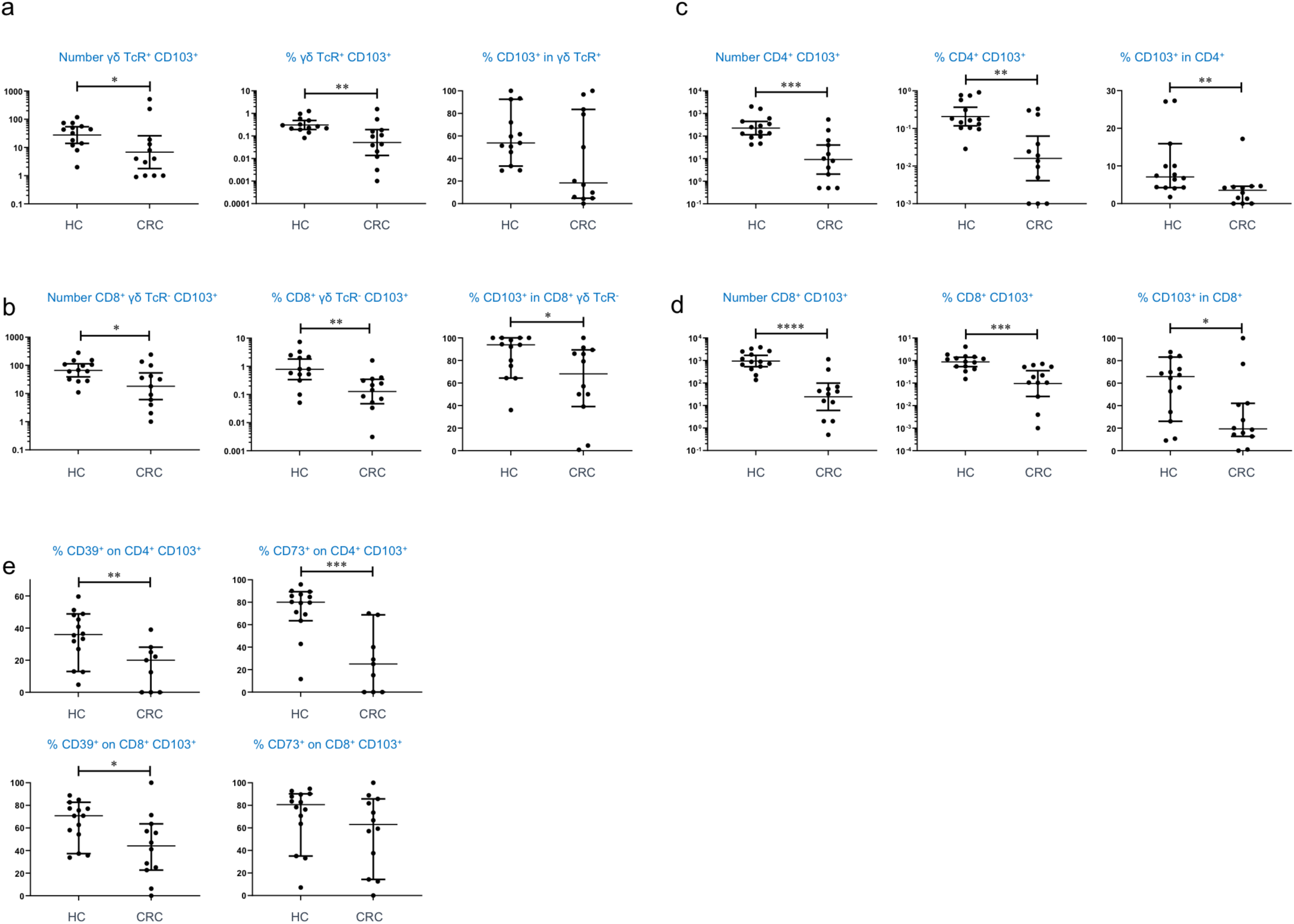
CRC is associated with depletion of γδ T-cells and Trm in colonic tissue and deficient regulatory function in CD4 Trm. A: Numbers and percentages of γδ CD103^+^ T-cells in IEL, alongside proportion of CD103 expression on total γδ T cells in healthy control (HC) and colorectal cancer (CRC) groups. B: Numbers and percentages of CD8^+^ γδ TCR^-^ CD103^+^ Trm in IEL, alongside proportions expressing CD103 of total CD8 T cells. C: Numbers and percentages of CD4^+^ CD103^+^ Trm in LPL, alongside proportions expressing CD103 of total CD4 T cells. D: Numbers and percentages of CD8^+^ CD103^+^ Trm in LPL, alongside proportions expressing CD103 of total CD8 T cells. E: Expression of regulatory T cell markers CD39 and CD73 on CD4 and CD8 Trm in LPL. HC: healthy controls, n=13; CRC: colorectal cancer, n=12. Median values ± 95% confidence intervals are shown; statistically significant differences between groups (Mann Whitney tests) are indicated.

We also analyzed expression of regulatory T cell markers on tissue resident cells (Figure 2e). We included CD73 as well as CD39, as it is also involved in extracellular ATP degradation to adenosine and regulatory function^16^ but is not selectively expressed on Trm^12^. Amongst LPL, reduced proportions of CD4^+^ Trm cells expressing both CD39 and CD73 were found in CRC. A similar trend (though not significant for CD73), was apparent in CD8^+^ Trm. Due to the extremely low numbers of cells detected in CRC tissue, expression of these markers in IEL could not be accurately compared. The data suggest some loss of regulatory T cell function in CRC-susceptible mucosa, perhaps secondary to an altered microbiota.

### Antibody responses to mucosa-associated bacteria are unaltered in CRC

IEM released from colonic biopsies were assessed for levels of IgA and IgG coated microbes, to determine whether there is an upregulated immune response to the intestinal microbiota in CRC, as we have described in IBD^12^. Interestingly, there was no significant difference in levels of either IgA or IgG coating in CRC compared to healthy controls, although the total numbers of IEM were reduced (Figure 3). A subset of 4 CRC patients had abnormally high IgG, but not IgA coating of their IEM, perhaps suggesting damage to the intestinal barrier.

**Figure 3.**
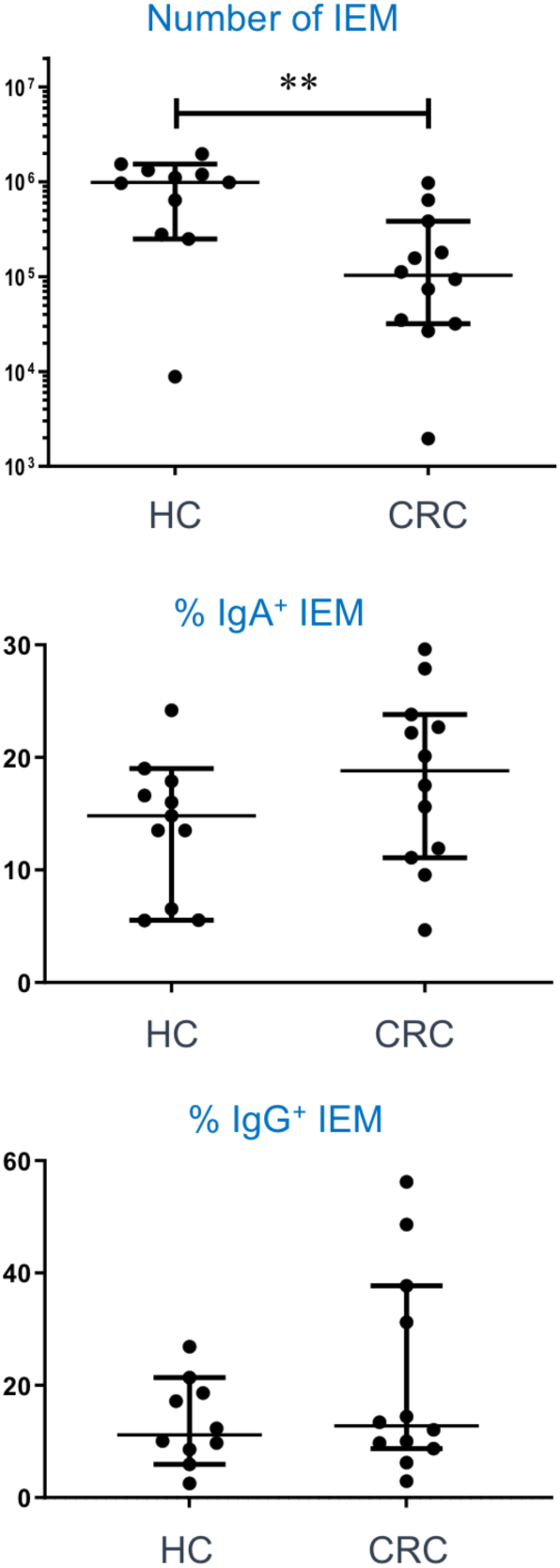
Antibody responses to mucosal-associated microbes are unchanged in CRC patients. IgA and IgG coating levels of IEM obtained from colonic biopsies of HC and CRC donors, after gating on SYBR Green^+^ events are shown. Upper graph: numbers of microbes recovered; middle graph: IgA coating; lower graph: IgG coating. Median values ± 95% confidence intervals are shown; statistically significant differences between groups (Mann Whitney tests) are indicated.

### T cell memory to commensal bacteria and fungi is perturbed while B cell memory is increased in CRC

To further assess microbiota:immune system interactions in CRC, we examined circulating acquired memory responses to a panel of 9 commensal intestinal (upper gut) bacteria and two intestinal yeast strains, using PBMC from CRC donors and healthy controls (HC). The bacterial strains were selected from a previously described panel based on their relatively high immunogenicity in an initial panel of 19^12^. Patterns of positive T cell and B cell proliferative responses to each strain showed a large degree of donor variability in both HC and CRC, with as expected, CD4^+^ T cell responses predominating (Figure 4). The overall levels of proliferation in CD4^+^ T cells showed a statistically significant difference between HC and CRC, although none of the comparisons for individual species were significant. The results were similar for CD8^+^ T cells, except that CD8 memory to *Staphylococcus epidermidis* was close to being significantly weaker in CRC than HC. By contrast, a dramatic difference was seen in B cell proliferative responses, with significantly greater B cell immunity to 5 of the 9 bacterial strains and also to *Candida albicans*, a yeast commonly found in the gut, in CRC. Overall levels of B cell response showed a statistically significant difference between HC and CRC, reflecting dysregulated B cell memory to both commensal bacteria and yeast.

**Figure 4.**
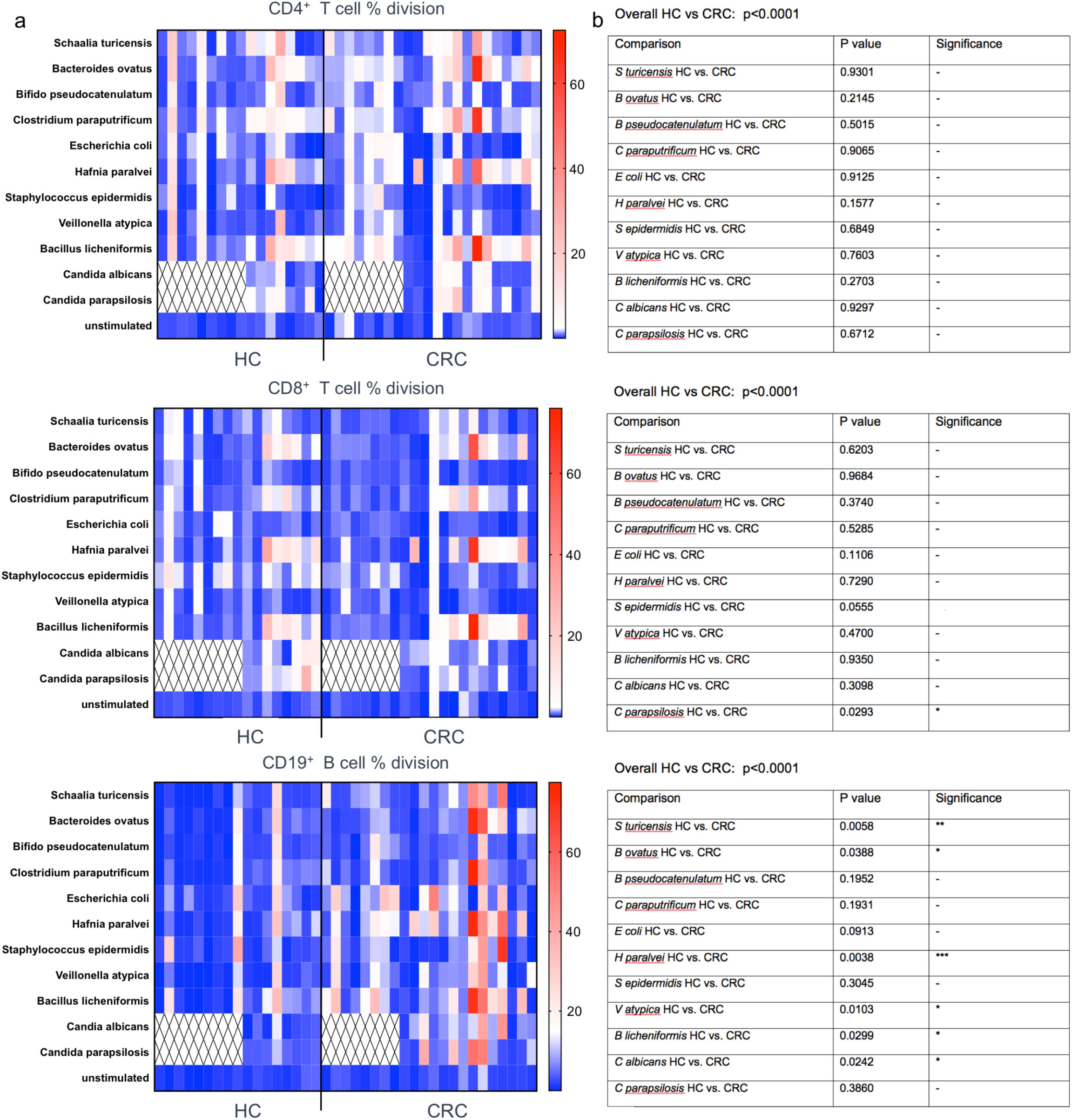
T- and B-cell memory responses to commensal bacteria and intestinal fungi are dysregulated in CRC. A: Heatmaps showing CD4/CD8 T-cell and B cell memory responses to indicated species, obtained from CellTrace Violet dilution data in gated populations of 7-day PBMC cultures with killed antigens. Each column shows results from one donor, with HC donors to the left and CRC donors to the right of the line. Numbers on scalebar represent % division. Hatched area: assays not performed. B: Corresponding statistical analysis of data in A, showing overall p value for variance within each subset and comparisons of responses for each individual species below. Kruskal-Wallis ANOVA was used to analyze data with correction for multiple comparisons applied using the Benjamini, Krieger & Yekutieli method; n=17 HC, n=22 CRC.

### Circulating transitional B cells, plasmablasts and effector memory B cells are increased in CRC

Since B cell memory to microbiota appeared dysregulated in CRC, we performed a global analysis of peripheral B cell subsets to further define perturbations of B cell homeostasis as potential biomarkers of CRC. We stained PBMC for naïve vs memory and effector memory B cell subsets, along with IgA- and IgG-switched B cells, plasmablasts and transitional B cells (T1 and T2). The data (Figure 5a and b) showed that overall proportions of memory B cells were not different between HC and CRC PBMC. However, proportions of effector memory B cells and plasmablasts (the most activated subsets) were significantly higher in CRC than HC. T1 and T2 transitional B cells, which are thought to be immature naïve B cells with high levels of autoreactivity^17^, were both found at higher levels in CRC than HC. There was no evidence of increased B cell switching to IgA or IgG from IgM. The activated signature of peripheral B cells in CRC led us to speculate whether they might contribute to the inflammation reported in CRC and other cancers^3,18^. We found that B cells from healthy donors activated overnight with PMA and ionomycin were indeed capable of producing both IFN-γ and TNF-α (Figure 5c), two pro-inflammatory cytokines present at higher levels in the circulation of CRC patients^3^.

**Figure 5.**
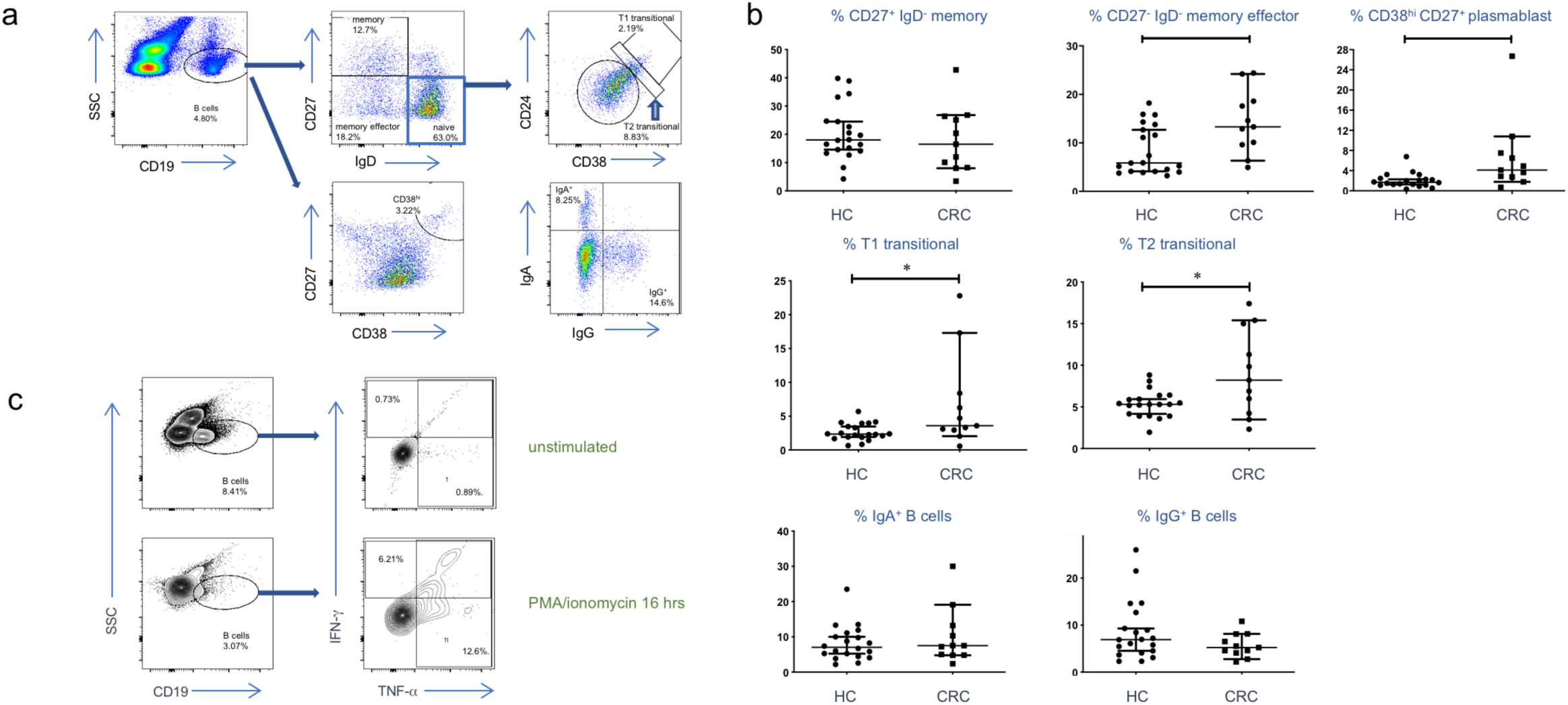
B-cell dysregulation is a systemic feature of CRC. A: Example of flow cytometric analysis of B cell subsets in PBMC. CD19^+^ B cells were allocated to naïve, memory and memory effector subsets based on IgD and CD27. Naive B cells were divided into T1 and T2 transitional B cells based on CD38 and CD24 expression. Total B cells were assessed for plasmablasts (CD38^hi^ CD27^+^ CD19^+^) and switching to IgA and IgG isotypes. B: Proportions of above subsets in HC vs CRC donor blood. Mann Whitney tests were used to compare groups (n=20 HC; n=11 CRC) and statistically significant differences are indicated. C: Intracellular cytokine staining of B cells from HC PBMC before and after stimulation with PMA and ionomycin, showing their capacity for TNF-α and IFN-γ production (representative of three independent experiments).

## Discussion

Identification of novel immune markers in tissue or blood that associate with “poor gut health” and susceptibility to CRC is of potential clinical value in prophylaxis and therapy. We describe an immune profile in CRC distinct from that in IBD that could be of use to identify IBD patients at highest risk of CRC.

A goal of tumor immunotherapy is to accelerate the generation of T cells with specificities capable of eliminating tumor cells, by blocking natural checkpoint inhibition pathways which prevent development of effector T cells with self-specificity. However, it is appreciated that there is a background level of anti-tumor immunity present in tissue, reflected by the large numbers of tissue resident lymphocytes in mucosal tissues at high risk of malignant transformation, such as the colon. γδ T cells have long been known to reside mainly in peripheral tissues and have anti-tumorigenic effects through unconventional specificities^19^. It appears that γδ T cells are the most effective cell type in tumor eradication^20^, due to their ability to recognize self-antigens whilst also retaining a degree of receptor variability. Their tissue-sensing capacity means that they are not dependent on the high mutational loads needed to generate peptide neoantigens for tumor recognition by conventional T cells^19^. Additionally, γδ T cells can respond to endogenous sterol intermediates which are expressed in malignant and virus-infected cells^21^. Our finding that these cells are deficient in CRC is therefore significant. The factors that sustain γδ T cells within specific tissues are unclear. However, it has been shown in mice that butyrophilin expression on intestinal epithelial cells is involved in the development of γδ IEL^22^, pointing towards interactions between epithelium and γδ T cells that could be dysregulated in CRC. Retinoic acid, an important mediator of T cell recruitment to the intestinal mucosa^23^, is deficient in colitis-associated CRC^24^ and also enhances IL-22 secretion from γδ T cells, which suppresses intestinal inflammation^25^. Furthermore, γδ T cell migration around intestinal villi is able to block translocation of pathogenic microbes^26^. Further research is required to determine how IEL populations can be boosted in order to maintain health^27^.

More recently, the definition of Trm as a distinct subset, residing for long periods in the epithelium and lamina propria of the colon, has expanded the repertoire of potential tissue-protective cells during homeostasis. The role of the intestinal microbiota, however, in the etiology of CRC is only beginning to become recognized^28^. The interplay between resident lymphocytes and the microbiota is the dominant feature of the colonic epithelium where tumors develop, and CRC has now been added to the growing list of diseases associated with an abnormal intestinal microbiome^29^. Moreover, developing tumors harbor their own microbiota^30^ and the Trm within tumors are associated with positive outcomes, suggesting that these cells can either prevent tumorigenesis or can expand in response to tumor- or microbiota-associated antigens during the course of the disease and kill target cells^31^.

The profound depletion of γδ T cells and Trm in the mucosa of CRC patients that we describe here suggests that there is an inherent susceptibility to tumorigenesis in these patients, linked to environmental or microbiota-related factors. Most patients were experiencing primary disease and had not received treatment with immunosuppressive agents or other medications that may have confounded the data, with the exception of pre-surgical antibiotic therapy. However, the latter was administered within the 24 hours prior to surgery so may not have impacted mucosal populations. Our observations that a similar paucity of IEL is seen in IBD patients, who are at increased risk of CRC, suggests that maintaining populations of γδ T cells and Trm at levels in seen healthy individuals is essential for maintaining homeostasis, blocking cell transformation and preventing inappropriate inflammation. Importantly, the expression of regulatory T cell molecules CD39 and CD73 on γδ T cells and Trm, including CD4 Trm in LPL, was reduced in CRC patients, strengthening the inference that immune barrier function and homeostasis are compromised in those developing CRC. The altered phenotype of mucosal CD4 T cells may also contribute to the dysregulation in B cell responses, since these depend on CD4 T cell “help”, balanced by CD4 regulatory T cell activity. γδ T cells are also known to have regulatory function which may balance their potent cytotoxic functions when present in tumor tissue^32,33^. These regulatory activities may suppress microbiota-dependent inflammation through the degradation of extracellular ATP released by microbes and activated immunocytes^34,35,36^, which we speculate also protects from parainflammation.

Impaired immune barrier function has been described as a consequence of aging or inflammation and is associated with CRC^37^. Although inflammation seen in CRC patients is not accompanied by increased secretory IgA synthesis against mucosa-associated microbes, we detected a strong B cell activation signal in CRC that was comparable to that seen in IBD. It is therefore possible that there is increased translocation or sampling of microbes by DC in the mucosa prior to tumorigenesis, perhaps triggered by an altered microbiota with expanded pathobiont species. Increased access of microbial antigens to mucosa-associated lymphoid tissue and mesenteric lymph nodes could have resulted in the increased B cell memory responses to a variety of commensal species, as well as the global activation of circulating B cells that we describe in CRC. Increased memory B cells and plasmablasts have previously been reported in CRC and shown to be present in tumor infiltrates^38^. Furthermore, the ability of B cells to secrete pro-inflammatory cytokines implicates them as a direct mediator of parainflammation, although there are numerous other potential cellular sources of these cytokines which have not been identified^39^.

In addition to the effector memory B cells and plasmablasts, which we identified as biomarkers in the blood, we also demonstrated increased proportions of transitional B cells in CRC patients. Transitional B cells are recent emigrants from bone marrow and are enriched in autoreactive cells^40^. Furthermore, intestinal tissue has been proposed as a site to which transitional B cells migrate and are selected to remove autospecificities, a process that is defective in systemic lupus erythematosus leading to uncontrolled autoantibody production^41^. Autoantibodies to tumor antigens are a known early indicator of CRC^42^. Therefore, perturbation of B cell repertoire selection by low-level barrier dysfunction in early CRC, as suggested by our data, may prevent negative selection of B cells specific for autoantigens and expand B cell clones reactive to microbiota-derived antigens. This, to our knowledge, is the first study to identify transitional B cell involvement in a non-B cell malignancy. It may point towards a common mechanism leading to loss of B cell tolerance towards both tumor-associated and microbial antigens during CRC etiology. This area clearly requires further investigation and mechanistic data acquired from animal models. Interestingly, in CRC the IgA responses to epithelia-associated microbes were unchanged, a result that contrasts with that in IBD patients^12^. Therefore, more overt mucosal inflammatory signals may be required before the secretory IgA pathway is disturbed.

Overall, our novel observations of altered microbiota:immune system interaction in CRC suggest that depleted gut-resident immune cells in genetically susceptible individuals may drive parainflammation and compromise immune surveillance in the intestinal mucosa, providing new insights into disease etiology. The activated B cell signature we have identified in CRC may prove useful in identifying high-risk individuals, while the profound deficiency in IEL emphasizes the importance of strategies that boost intestinal cellular immunity for prophylaxis. Our data also strengthen the rationale for developing γδ T cell therapy for treatment of cancer^43^.

## Data Availability

The data that support the findings of this study are available from, upon reasonable request.

## Abbreviations

CRC: colorectal cancer
Trm: resident memory T-cells
HC: healthy control
IBD: inflammatory bowel disease
IEL: intraepithelial lymphocytes
LPL: lamina propria lymphocytes
IEM: intraepithelial microbes
TNF: tumor necrosis factor
IFN: interferon
DC: dendritic cell

## Acknowledgements

We thank Alison Scoggins for administrative assistance. We are grateful to the staff and patients of St Mark’s Hospital and our blood donors for their participation in this study.

