## Supplemental Table 1 for "Altered immunity to microbiota, B cell activation and depleted γδ / resident memory T cells in colorectal cancer"

Supplementary Table 1. Antibodies used:

| **ANTIGEN** | **FLUOROCHROME** | **mAb CLONE** | **SUPPLIER** |
|---|---|---|---|
| CD4 | BB515 | SK3 | BD Biosciences |
| CD8α | BV605 | SK1 | BioLegend |
| CD19 | PE-Cy5 | HIB19 | BD Biosciences |
| CD24 | APC-Cy7 | ML5 | BioLegend |
| CD27 | PE | M-T271 | BD Biosciences |
| CD38 | BV605 | HIT2 | BioLegend |
| CD39 | PE-Cy7 | A1 | BioLegend |
| CD69 | BV605 | FN50 | BioLegend |
| CD73 | APC | AD2 | BioLegend |
| CD103 (integrin αE) | PE | Ber-ACT8 | BioLegend |
| IgA | APC | IS11-8E10 | Miltenyi Biotec |
| IgD | PE-Cy7 | IA6-2 | BD Biosciences |
| IgG | FITC | IS11-3B2.2.3 | Miltenyi Biotec |
| IgM | BV421 | G20-127 | BD Biosciences |
| Integrin β7 | PE | FIB504 | BD Biosciences |
| IFN-γ | FITC | B27 | BD Biosciences |
| Runx3 | PE | R3-5G4 | BD Biosciences |
| TCR γδ | FITC | 11F2 | BD Biosciences |
| TNF-α | PE-Cy7 | MAb11 | eBioscience |
